# “The injection gives freedom” - An exploration of long-acting injectable HIV treatment acceptance among patients seeking care in two Nairobi tertiary hospitals

**DOI:** 10.1101/2025.02.26.25322923

**Authors:** Nyukuri Duncan Wekesa, Abongo Melanie, Anastacia Mbithi, Christine Kundu, Mutai Kenneth, Reena Shah

## Abstract

**Background:** Long-acting injectable antiretroviral therapy offers an alternative to daily oral HIV treatment, potentially improving adherence and reducing stigma. While its adoption has been successful in some settings, evidence on its acceptability in sub-Saharan Africa, outside clinical trials, remains limited.

**Methods:** We conducted a mixed-methods study among people living with HIV (PLHIV) in two tertiary hospitals in Nairobi, Kenya—a public facility (Kenyatta National Hospital) and a private facility (Aga Khan University Hospital). A cross-sectional survey of 356 participants assessed awareness, willingness to switch, and predictors of acceptability. In addition, 30 participants took part in three focus group discussions (FGDs) exploring perceptions, concerns, and system-level considerations. Quantitative data were analyzed using descriptive and inferential statistics, while qualitative data were analyzed thematically.

**Results:** Overall, 72.2% of survey participants indicated willingness to switch to LAI-ART. Prior awareness of LAI-ART was the strongest independent predictor of acceptability (aOR 4.03, 95% CI: (1.45-11.18). Drivers of interest included reducing pill burden (77.8%), improving adherence (55.6%), and maintaining privacy (45.5%). Despite high acceptability, FGDs revealed cautious optimism, with concerns about side effects, treatment rigidity, and logistical challenges such as travel and clinic access.

**Conclusion:** These findings suggest that while LAI-ART is seen as promising, its uptake will depend on education, trust-building, and reliable delivery systems. Successful introduction in Kenya will require patient-centered communication, system readiness, and equitable access strategies.

## Introduction

Long-acting injectable antiretroviral therapy (LAI-ART) offers a promising alternative to the daily pill regimen for treating HIV. Administered every two months, this innovative therapy could simplify treatment, improve patient adherence, and boost retention in care (1,2).

This is especially critical in sub-Saharan Africa (SSA), a region that carries two-thirds of the 40 million people living with HIV worldwide (3). In Kenya, for example, where 1.4 million people are living with HIV, persistent adherence challenges are common due to factors such as pill burden, treatment fatigue, and stigma (4). These issues are particularly pronounced among adolescents and young people, who often have significantly lower adherence rates than older adults due to heightened stigma and challenges during their transition to adulthood (5). Consequently, there is an urgent need to explore new and creative strategies to address the unique challenges of the region.

Studies in high-income countries have shown that LAI-ART is as effective as daily oral therapy at maintaining viral suppression, and patient satisfaction is high (6–12). In the US-based CUSTOMIZE implementation study, participants reported that 92% preferred injectable therapy over their previous oral regimen, and 97% indicated willingness to continue long-acting injections(13).

While there is compelling evidence from North America and Western Europe, there are significant data gaps in sub-Saharan Africa. A 2024 study in Uganda by Zakumumpa et found that 94% of participants in a demonstration project wanted to continue with LAI-ART(14).

However, the results may be biased since participants were already on the treatment. A South African study showed that while only 12% of adolescents and young people with HIV initially preferred LAI-ART, this preference increased to 66% among those who faced multiple adherence challenges like stigma and pill burden(15). Research from the Dominican Republic, Tanzania, and a small 2018 study in Kenya also showed broad acceptance of LAI-ART, driven by the desire for reduced stigma and greater privacy (16). A recent scoping review also found high acceptability for LAI-ART in sub-Saharan Africa, ranging from 63% to 98%(17). Still, most of these studies were conducted among specific populations, like those in clinical trials or high-risk, HIV-negative individuals, which limits their broader applicability.

This study builds on prior work by examining the acceptability of LAI-ART among a more general population of PLHIV receiving care in two tertiary hospitals in Nairobi—one public and one private. Using a mixed-methods approach grounded in the Consolidated Framework for Implementation Research (CFIR)(18), we explore perspectives of patients currently on oral ART who have not yet used injectables. By addressing both individual and system-level factors, this study provides actionable insights to support equitable LAI-ART implementation in Kenya and similar settings across SSA.

## Materials and methods

### Study design

This was an exploratory mixed-methods study using a convergent parallel design, conducted to assess the acceptability of long-acting injectable antiretroviral therapy among people living with HIV in Kenya. Quantitative data was gathered through a survey questionnaire administered to the patients while qualitative data was obtained through three focus group discussions. Quantitative and qualitative dada were analyzed independently before being integrated during interpretation. Importantly, this was not a clinical trial; participants were not enrolled into any intervention arm, and no medications were administered. The Consolidated Framework for Implementation Research (CFIR) guided the development of survey tools and focus group guides, and later informed the interpretation of qualitative themes to explore multi-level barriers and facilitators to LAI-ART adoption (18).

### Study site and participants

The study was conducted at two tertiary hospitals in Nairobi, Kenya: Aga Khan University Hospital (AKUH) and Kenyatta National Hospital (KNH). AKUH, a private not-for-profit institution, is a referral center offering specialized care. KNH is the largest public hospital in Kenya and a teaching facility for the University of Nairobi, supporting over 8,000 PLHIV in outpatient care.

The study was conducted at AKUH between December 14, 2023, and January 12, 2024, and at KNH between May 21, 2024, and July 9, 2024. Eligible participants were adults (≥18 years), currently receiving oral ART, and enrolled in HIV outpatient care at either facility. Participants were excluded if they had cognitive impairments, were acutely unwell, or could not communicate in English or Kiswahili.

### Sampling and recruitment

Quantitative participants were selected using systematic random sampling from outpatient clinic attendance logs to achieve a representative sample. The sample size was calculated using Cochran’s formula for cross sectional studies(19) with a 95% confidence level and a 5% margin of error. The acceptability rate of 66%, which was used for the calculation, was derived from a South African study by Toska et al(15). This study explored the acceptability of long-acting injectable LAI-ART among adolescents and young people living with HIV (AYLHIV) who were currently on oral therapy, and this prevalence was chosen because it yielded the largest sample size. This resulted in a minimum required sample size of 345 participants.

Participants who consented to participate in the survey were approached during clinic visits and invited to take part in focus group discussions. The participants were selected via purposive sampling based on gender, age, and clinic site to capture diverse perspectives.

### Data collection procedures

A structured questionnaire was used to collect data on demographics, knowledge of LAI-ART, and attitudes toward injectable regimens. The data collection tool used had been developed and validated by Simoni et al(20) in a similar study conducted in Tanzania and Dominican and it was also based on CFIR constructs. Data were collected in person or via REDCap online survey links.

### Focus group discussions (FGDs)

Three FGDs were conducted, with 10 participants per FGD – the recommended number consisting of between 6 and 12 participants per group(21) – to foster open discussion.

Discussions were held in private rooms at each study site and moderated by M.A, an experienced qualitative researcher in conducting FGDs, with no clinical relationship to participants. A semi-structured discussion guide covered themes such as LAI-ART awareness, perceived benefits and drawbacks, implementation concerns, and facility readiness. Sessions were audio-recorded with consent; field notes were not taken. Discussions continued until thematic saturation was reached. Thematic saturation was assessed by the point at which no new themes emerged during subsequent discussions

### Data analysis

#### Quantitative analysis

Survey data were analyzed using SPSS version 25. Descriptive and inferential analyses were performed. Participant characteristics were summarized by study site (KNH and AKUH) and for the combined sample. Categorical variables, including demographic characteristics, social history, and medical history, were presented as frequencies and percentages.

Bivariate Analysis: Comparisons between KNH and AKUH participants were conducted using the Chi-square test of independence; Fisher’s exact test was applied when expected cell counts were fewer than five.

Multivariable Analysis: The primary outcome was acceptability of LAI-ART (Yes vs No). Predictors of acceptability were assessed using logistic regression. Bivariate logistic regression was first used to estimate crude odds ratios (OR) with 95% confidence intervals (CI). Multivariate logistic regression included study site and all key demographic, social, and awareness-related variables to estimate adjusted odds ratios (aOR). Statistical significance was set at p < 0.05.

#### Qualitative analysis

Audio recordings from focus group discussions were transcribed verbatim. For each transcript, structured summary templates were completed using pre-specified domains derived from the CFIR (S1 Fig). Two independent coders (DN and CK) reviewed each transcript to identify blocks of text corresponding to relevant CFIR constructs. Data were aggregated across participants to facilitate comparison within and across domains. The two researchers discussed coding decisions, resolved discrepancies through consensus, and agreed on emergent themes. We did not return transcripts to participants for feedback or validation due to logistical and confidentiality constraints. However, the use of a structured summary template, independent dual coding, and consensus discussions enhanced the reliability and rigor of the analysis.

#### Integration of findings

Quantitative and qualitative data were integrated during interpretation through triangulation, identifying areas of convergence and divergence. Results were jointly displayed to enhance understanding of complex, multi-level factors influencing LAI-ART acceptability.

### Ethical Statement

Ethical approval for this study was obtained from both the Kenyatta National Hospital– University of Nairobi Ethics and Research Committee (Approval No. P816/10/2023) and the Aga Khan University Institutional Research and Ethics Committee (Approval No. 2023/ISERC-50(v3)). All participants provided written informed consent prior to enrollment, after receiving detailed explanations of the study’s objectives, procedures, potential risks, and measures taken to protect confidentiality.

Participation was entirely voluntary, and refusal or withdrawal did not affect participants’ access to clinical care. Data were anonymized at the point of collection, and no personal identifiers were linked to survey or focus group responses. Focus group discussions were conducted in private rooms to safeguard participants’ privacy and minimize stigma-related risks. A trained, experienced and independent moderator facilitated the discussions in a manner sensitive to participants’ experiences. All findings are reported in aggregate to ensure that no individual can be identified.

## Results

A total of 356 participants were enrolled, with the majority from KNH (77.2%) (Table 1 and S1 Table). The cohort was predominantly female (59.0%), with the largest group aged 40–49 years (32.3%). Most participants were married (61.2%), employed (81.5%), and had children (81.7%). Nearly all were on once-daily ART (95.5%), with the vast majority describing their treatment as simple (91.6%) and well tolerated (86.0%). More than one-third had lived with HIV for over 10 years (35.4%), and common comorbidities included hypertension (16.6%), diabetes (6.5%), and high cholesterol (5.3%).

**Table 1.** Demographics and Social History.

| Characteristic | Category/Statistic | KNH<br>(N=275) | AKUH<br>(N=81) | Combined<br>(N=356) | p-value<br>(unadjusted) |
| --- | --- | --- | --- | --- | --- |
| **Age group** | 20-29 | 29 (10.5%) | 2 (2.5%) | 31 (8.7%) | 0.0001 |
|  | 30-39 | 83 (30.2%) | 10 (12.3%) | 93 (26.1%) |  |
|  | 40-49 | 86 (31.3%) | 29 (35.8%) | 115 (32.3%) |  |
|  | 50-59 | 66 (24.0%) | 30 (37.0%) | 96 (27.0%) |  |
|  | 60-69 | 10 (3.6%) | 10 (12.3%) | 20 (5.6%) |  |
|  | 70-79 | 1 (0.4%) | 0 (0.0%) | 1 (0.3%) |  |
| **Gender** | Female | 174 (63.3%) | 36 (44.4%) | 210 (59.0%) | 0.0031 |
|  | Male | 101 (36.7%) | 45 (55.6%) | 146 (41.0%) |  |
| **Marital status** | Divorced | 3 (1.1%) | 4 (4.9%) | 7 (2.0%) | 0.061 |
|  | I prefer not to say. | 1 (0.4%) | 0 (0.0%) | 1 (0.3%) |  |
|  | Married | 168 (61.1%) | 50 (61.7%) | 218 (61.2%) |  |
|  | Separated | 12 (4.4%) | 8 (9.9%) | 20 (5.6%) |  |
|  | Single | 71 (25.8%) | 16 (19.8%) | 87 (24.4%) |  |
|  | Widow(er) | 20 (7.3%) | 3 (3.7%) | 23 (6.5%) |  |
| **Living arrangement** | Alone | 58 (21.1) | 29 (35.8) | 87 (24.4) | 0.0009 |
|  | As A Couple | 15 (5.5) | 10 (12.3) | 25 (7.0) |  |
|  | With Family | 202 (73.5) | 42 (51.9) | 244 (68.5) |  |
| **Has children** | No | 46 (16.7%) | 19 (23.5%) | 65 (18.3%) | 0.191 |
|  | Yes | 229 (83.3%) | 62 (76.5%) | 291 (81.7%) |  |
| **Employment status** | I prefer not to say | 2 (0.7%) | 0 (0.0%) | 2 (0.6%) | 0.737 |
|  | No | 49 (17.8%) | 14 (17.3%) | 63 (17.7%) |  |
|  | Yes | 224 (81.5%) | 66 (82.7%) | 290 (81.5%) |  |

Comparisons between sites revealed important differences. AKUH participants were more likely to be male (55.6% vs. 36.7%, p = 0.003), older (37.0% aged 50–59 vs. 24.0% at KNH, p < 0.001), and to report diabetes (14.8% vs. 4.0%, p = 0.001) and high cholesterol (12.3% vs. 3.3%, p = 0.004). They were also more likely to have lived with HIV for over 10 years (53.1% vs. 30.2%) and to have changed ART regimens (45.7% vs. 17.8%, p < 0.001), mainly due to tolerance issues (35.8% vs. 8.4%, p < 0.001). In contrast, KNH participants were more likely to be married (61.1% vs. 50.6%, p = 0.004), to live with family (73.5% vs. 51.9%, p = 0.001), and to report psychiatric disorders (11.8% vs. 2.5%). They also reported missing ART doses more often, with 58.2% acknowledging occasional missed doses compared to 21.0% at AKUH (p < 0.001). Patterns of care utilization differed as well: 91.4% of AKUH participants attended clinic every 2–3 months compared to 61.1% at KNH (p < 0.001).

### Knowledge and acceptability of lai-art

Overall, 65.2% of participants had prior knowledge of LAI-ART, most commonly from a healthcare provider (57.3%) or online resources (55.2%). Acceptability was high, with 72.2% expressing willingness to use LAI-ART. However, 17.4% reported they lacked sufficient information to decide, and 8.4% indicated they would not accept it. (Table 2)

**Table 2.** LAI-ART awareness & acceptability.

| Characteristic | Category/Statistic | KNH<br>(N=275) | AKUH<br>(N=81) | Combined<br>(N=356) | p-value<br>(unadjusted) |
| --- | --- | --- | --- | --- | --- |
| Aware of LAI-ART | No | 99 (36.0%) | 25 (30.9%) | 124 (34.8%) | 0.428 |
|  | Yes | 176 (64.0%) | 56 (69.1%) | 232 (65.2%) |  |
| Source of information | Healthcare provider | 102 (58.0) | 31 (55.4) | 133 (57.3) | 0.852 |
|  | Support group | 59 (33.5) | 11 (19.6) | 70 (30.2) | 0.071 |
|  | Online resources | 87 (49.4) | 41 (73.2) | 128 (55.2) | 0.003 |
|  | Other | 1 (0.6) | 2 (3.6) | 3 (1.3) | 0.145 |
| Acceptability of LAI-ART | Yes | 206 (74.9) | 51 (63.0) | 257 (72.2) | 0.002 |
|  | No | 25 (9.1) | 5 (6.2) | 30 (8.4) |  |
|  | I Don't have enough information | 42 (15.3) | 20 (24.7) | 62 (17.4) |  |
|  | I am not sure | 2 (0.7) | 5 (6.2) | 7 (2.0) |  |

Site differences were observed. Acceptability was higher at KNH (74.9%) compared to AKUH (63.0%, *p* = 0.002). KNH participants were more likely to learn about LAI-ART from healthcare providers, whereas AKUH participants more frequently cited online sources (*p* = 0.003).

### Perceived advantages and disadvantages of LAI-ART

Among those willing to use LAI-ART, the most frequently cited advantages were the ability to stop daily pill-taking (77.8%), to avoid forgetting doses (55.6%), and to reduce daily focus on HIV (50.2%) (S2 Table). Other benefits included improved privacy (45.5%) and the ability to “forget the disease” (37.0%).

The most common disadvantages were fear of side effects (68.1%), fear of injections (28.0%), and concerns about being treated as “guinea pigs” (17.9%) (S3 Table). Site differences emerged: AKUH participants were more likely to cite effectiveness and dose adherence as advantages, but also more frequently reported fear of side effects. In contrast, KNH participants more often valued privacy-related benefits and reported fear of injections.

### Preferences for dosing

In terms of dosing, the majority favored longer injection intervals, with three months (27.2%) and six months (35.8%) being the most common choices **(S4 Table)**. Preferences varied by site, with AKUH participants more likely to prefer two-monthly dosing, while KNH participants leaned toward six-monthly schedules.

### Predictors of acceptability of LAI-ART

Regression analysis identified prior awareness of LAI-ART as the strongest independent predictor of acceptability **(Table 3)**. Participants who had heard of LAI-ART were more than four times as likely to express willingness to use it compared to those unaware (AOR 4.03; 95% CI 1.45–11.18, *p* = 0.007). Other factors, including study site, marital status, and online sources of information, were not significant after adjustment.

**Table 3.** Predictors of LAI-ART Acceptability.

| Predictor | Crude OR (95% CI) | Adjusted OR (95% CI) | p-value |
| --- | --- | --- | --- |
| Study site | 0.57 (0.34-0.96) | - | 0.036 |
| Marital status | 0.36 (0.14-0.94) | - | 0.037 |
| Awareness of LAI-ART | 2.68 (1.66-4.32) | - | 0.0 |
| Learned from online resources | 1.98 (1.18-3.32) | - | 0.01 |
| Awareness of LAI-ART (Yes vs No) | - | 4.03 (1.45-11.18) | 0.007 |

### Qualitative findings and integrated analysis

A summary of emergent themes and sub-themes across CFIR domains, along with illustrative quotes, is presented in Table 4.

**Table 4.**
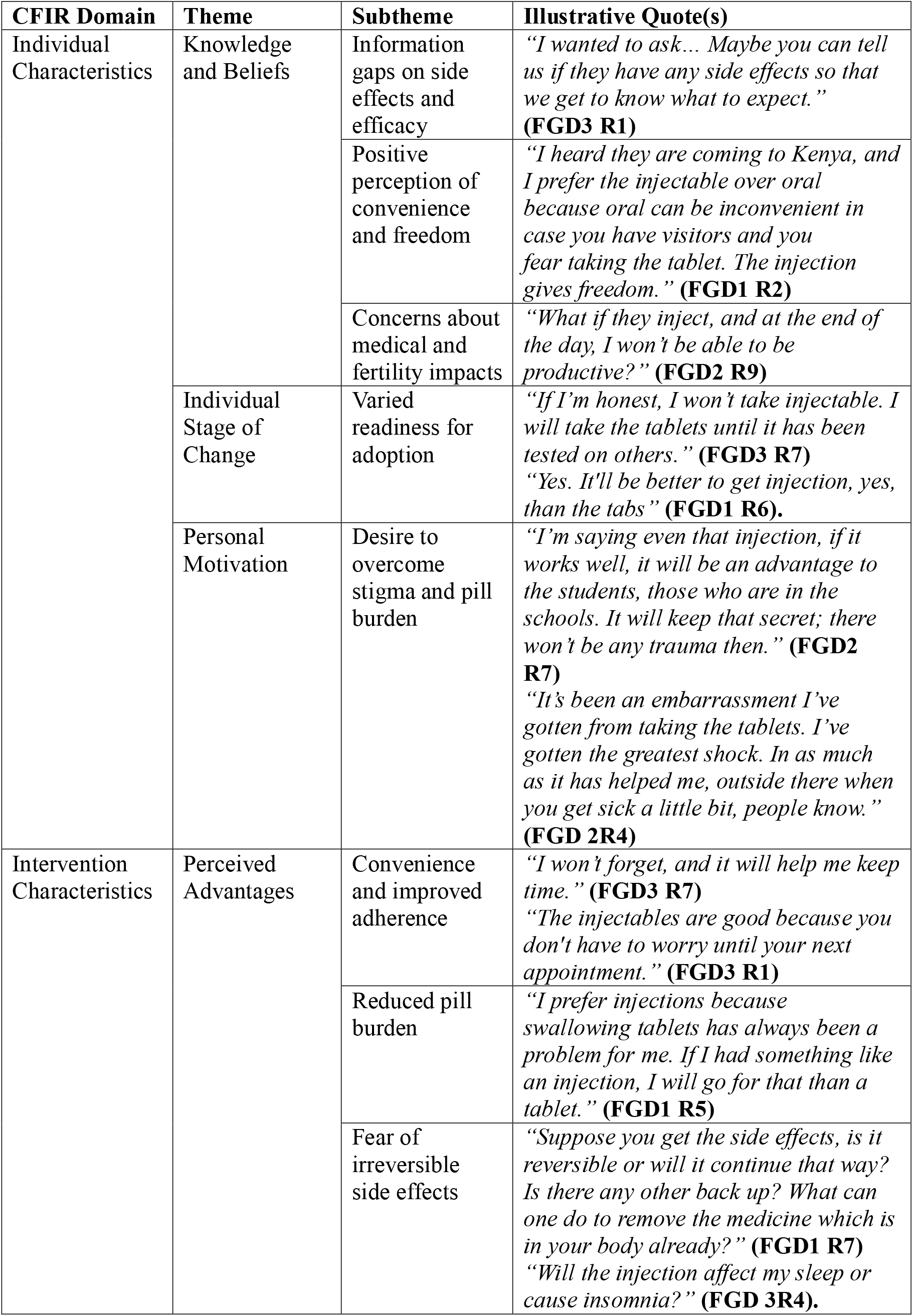
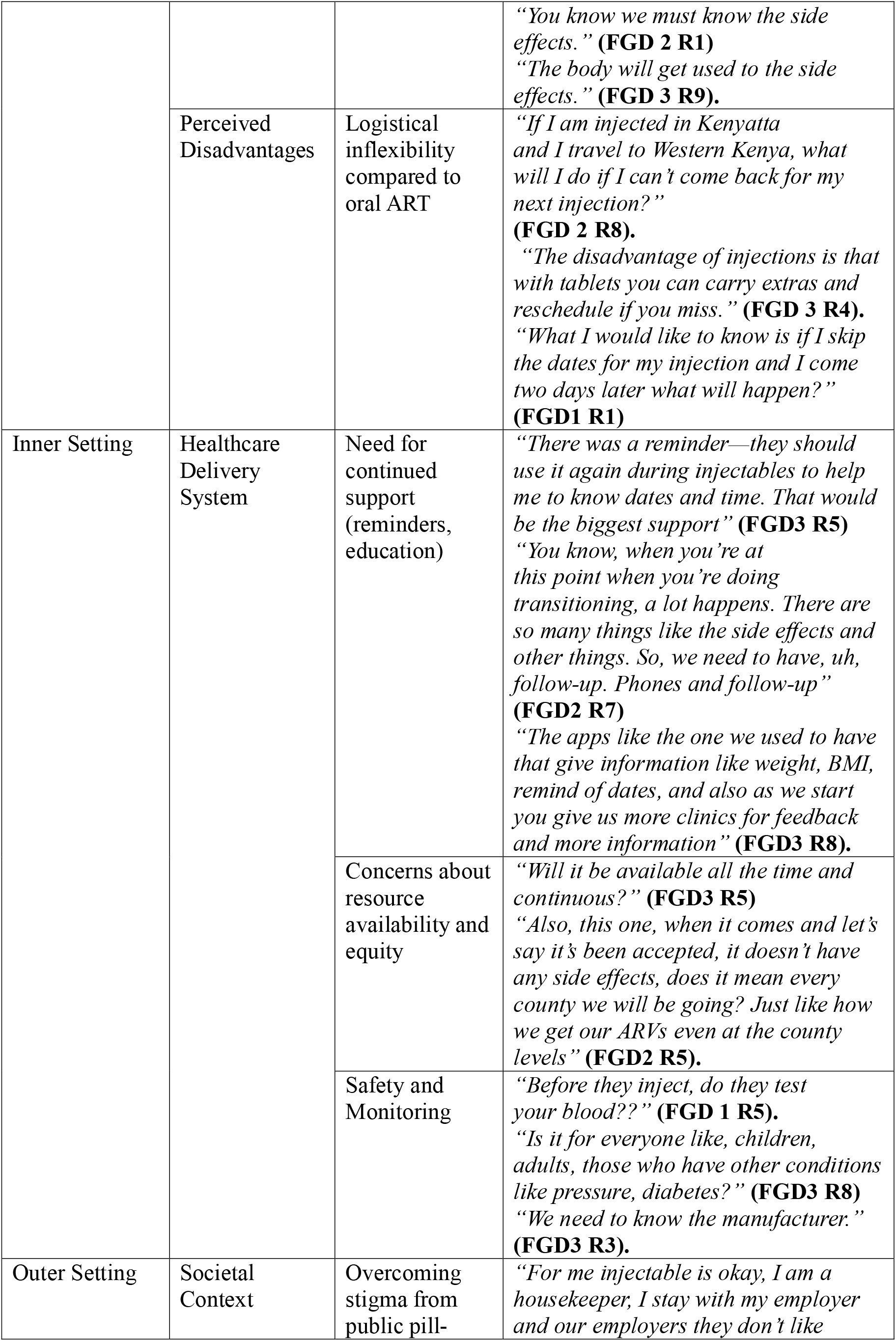
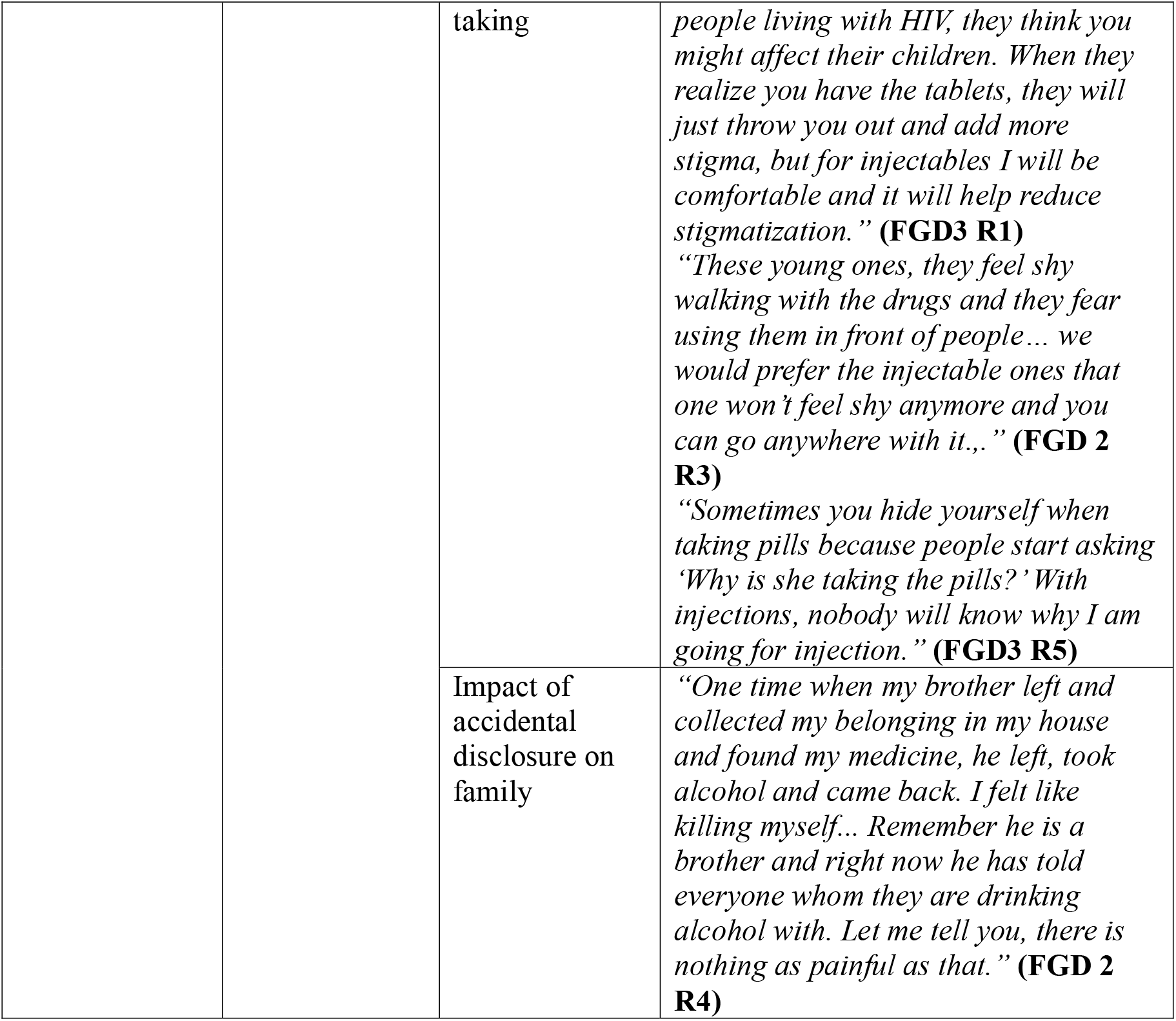
Emergent themes and sub-themes from qualitative analysis based on the CFIR.

### Convergence: Reinforcing survey data

#### The Burden of Pills and Stigma

Survey data showed limited disclosure of HIV status: only 12.4% had disclosed to friends and 6.3% to colleagues. Also, nearly half (45.5%) cited the ability to hide treatment for privacy as a perceived advantage of LAI-ART. FGDs contextualized this, with participants describing stigma and discretion as strong motivations:

> *“I prefer the injectable over oral because oral can be inconvenient in case you have visitors and you fear taking the tablet. The injection gives freedom*.*”* **(FGD1, R2)**
>
> *“For me injectable is okay, I am a housekeeper…our employers don’t like people living with HIV. When they realize you have the tablets, they will just throw you out and add more stigma, but for injectables I will be comfortable and it will help reduce stigmatization*.*”* **(FGD3, R1)**

#### Fear and Distrust of New Treatments

Quantitatively, 68.1% cited fear of side effects, 28.0% feared injections, and 17.9% worried about being treated as “guinea pigs.” FGDs elaborated that the main concern was the irreversibility of long-acting drugs and the desire to see them proven safe first:

> *“If it has already been injected into the body and it has to stay for 3 months but you are very reactive, it will be difficult to stop—for example, allergies*.*”* **(FGD2, R1)**
>
> *“If I’m honest, I won’t take injectable. I will take the tablets until it has been tested on others*.*”* **(FGD3, R7)**

#### Adherence Motivation

Survey findings showed that 55.6% of those willing to switch valued LAI-ART for helping to avoid missed doses. FGDs reinforced this, highlighting both practical and psychological benefits:

> *““I prefer injections because swallowing tablets has always been a problem for me. If I had something like an injection, I will go for that than a tablet*.*”* **(FGD 1R5.)**
>
> *“The injectables are good because you don’t have to worry until your next appointment*.*”* **(FGD 3 R1)**.

### Divergence and nuance: Highlighting tensions

#### Cautious Optimism vs. High Acceptability

Although surveys showed 72.2% acceptability, qualitative data revealed this represented a form of cautious optimism rather than unconditional support. Many participants preferred a trial period to assess safety before committing long-term:

> *“I will take the injection for three months to weigh the side effects, then commit for a year if it reacts positively*.*”* **(FGD3, R1)**

#### Convenience vs. Flexibility

Survey responses suggested broad willingness to use LAI-ART, but FGDs revealed concerns about reduced flexibility compared to oral ART, particularly when travel or missed appointments were involved:

> *“The disadvantage of injections is that with tablets you can carry extras and reschedule if you miss*.*”* **(FGD3, R4)**
>
> *“If I am injected in Kenyatta and I travel to Western Kenya, what will I do if I can’t come back for my next injection?”* **(FGD2, R8)**

#### System-Level Demands Beyond the Injection

FGDs highlighted broader health system challenges not reflected in survey data, including readiness for implementation, equitable access across counties, and the need for monitoring and follow-up:

> *“You know, when you’re at this point when you’re doing transitioning, a lot happens… So we need to have follow-up. Phones and follow-up*.*”* **(FGD2)**
>
> *“…does it mean every county we will be going? Just like how we get our ARVs even at the county levels?”* **(FGD2, R5)**
>
> *“Before they inject, do they test your blood?”* **(FGD1, R5)**

## Discussion

This mixed-methods study found that long-acting injectable ART (LAI-ART) was highly acceptable among people living with HIV in Nairobi, with over 70% expressing willingness to switch from oral therapy. Acceptability was driven by relief from pill burden, stigma reduction, and improved adherence, while concerns about side effects, injections, and system-level readiness tempered enthusiasm. Prior awareness of LAI-ART emerged as the strongest independent predictor of acceptability, suggesting that education and communication will be pivotal to uptake. Importantly, willingness to adopt was not unconditional but reflected a cautious optimism, with participants calling for a trial period before long-term commitment.

Our results align with evidence from other sub-Saharan African settings while also contributing novel insights(14–16,22,23). In Uganda, acceptability exceeded 90% among participants already enrolled in demonstration projects for LAI-ART, though this likely reflects selection bias since patients had firsthand experience with the regimen. Toska et al in South African study among adolescents and young adults reported much lower baseline preference (~12%), but willingness increased markedly among those facing multiple adherence barriers, illustrating the contextual nature of acceptability(15). By contrast, our study examined a broader population of PLHIV in routine care across both public and private hospitals, showing that interest is substantial even before introduction. Differences observed across sites likely reflect underlying sociodemographic and structural contexts. For example, higher acceptability at KNH may be explained by greater exposure to HIV-related stigma and higher rates of missed oral doses, making the privacy and adherence benefits of LAI-ART more appealing. In contrast, AKUH participants—who reported longer HIV duration, more regimen changes, and greater access to online information—expressed more reservations, possibly reflecting higher health literacy, prior treatment side effects, and greater access to diverse information sources, including negative portrayals. These contextual variations emphasize that acceptability is shaped not only by biomedical efficacy but also by patients lived experiences and care environments.

Beyond individual preferences, participants emphasized system-level considerations. Concerns about reliable supply chains, continuity of treatment during travel, and decentralization beyond tertiary hospitals highlight the importance of ensuring readiness within Kenya’s mixed health system. Without attention to these structural elements, the psychosocial and adherence benefits of LAI-ART risk being undermined.

## Strengths and limitations

This study is among the first mixed-methods analyses of LAI-ART acceptability in Kenya, combining quantitative and qualitative data from both public and private tertiary facilities. The relatively large sample strengthens generalizability to similar urban populations, and the qualitative data enrich interpretation of survey findings. However, the study was limited to Nairobi, and findings may not represent rural or lower-level facilities where system constraints differ. Awareness of LAI-ART was moderate at 64%, and reported willingness may not fully predict future uptake once available. Social desirability bias cannot be excluded, and as a cross-sectional study, we cannot capture how attitudes might evolve with greater exposure or rollout.

## Conclusion

In summary, while LAI-ART is highly acceptable, uptake will depend on raising awareness, addressing stigma, building trust, and ensuring system preparedness. Tailoring implementation to different patient populations and facility contexts will be key to realizing its promise as a transformative option for HIV care in sub-Saharan Africa.

## Data Availability

All data produced in the present study are available upon reasonable request to the authors

## Supporting information

**S1 Table**. Medical & HIV/ART History

**S2 Table**. Advantages of LAI-ART among responders

**S3 Table**. Disadvantages of LAI-ART among responders

**S4 Table**. Preferences for Dosing

**S1 Fig**. Conceptual Framework for Acceptability of LAI-ART using the Consolidated Framework for Implementation Research

