## Supplementary Figure 1 for "“The injection gives freedom” - An exploration of long-acting injectable HIV treatment acceptance among patients seeking care in two Nairobi tertiary hospitals": S1 _ Fig.docx

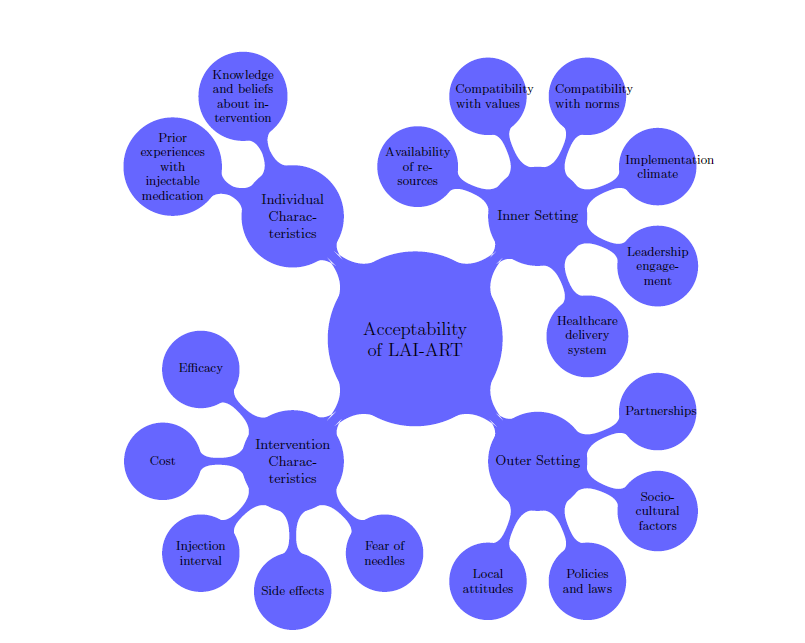

S1 Fig. Conceptual Framework for Acceptability of LAI-ART using the Consolidated Framework for Implementation Research
