## Supplementary Table 1 for "“The injection gives freedom” - An exploration of long-acting injectable HIV treatment acceptance among patients seeking care in two Nairobi tertiary hospitals": S1 _ Table.docx

**S1 Table. Medical & HIV/ART History**

| **Characteristic** | **Category/Statistic** | **KNH (N=275)** | **AKUH (N=81)** | **Combined (N=356)** | **p-value** |
| --- | --- | --- | --- | --- | --- |
| Diabetes | N (%) | 11 (4.0) | 12 (14.8) | 23 (6.5) | **0.001**. |
| High blood pressure | N (%) | 42 (15.3) | 17 (21.0) | 59 (16.6) | 0.296 |
| High cholesterol | N (%) | 9 (3.3) | 10 (12.3) | 19 (5.3) | 0.004 |
| Psychiatric disorder/depression | N (%) | 32 (11.8) | 2 (2.5) | 34 (9.6) | 0.032 |
| Other medical history | N (%) | 34 (12.4) | 9 (11.2) | 43 (12.1) |  |
| Duration since HIV infection | <1 year | 20 (7.3%) | 7 (8.6%) | 27 (7.6%) | <0.001 |
|  | 1–5 years | 80 (29.1%) | 14 (17.3%) | 94 (26.4%) |  |
|  | 5–10 years | 62 (22.5%) | 17 (21.0%) | 79 (22.2%) |  |
|  | >10 years | 83 (30.2%) | 43 (53.1%) | 126 (35.4%) |  |
|  | Prefer not to say | 30 (10.9%) | 0 (0.0%) | 30 (8.4%) |  |
| Currently taking ART | More than twice/day | 1 (0.4%) | 0 (0.0%) | 1 (0.3%) | 0.855 |
|  | Once/day | 262 (95.3%) | 78 (96.3%) | 340 (95.5%) |  |
|  | Twice/day | 11 (4.0%) | 3 (3.7%) | 14 (3.9%) |  |
| ART treatment perception | Complicated | 23 (8.4%) | 6 (7.4%) | 29 (8.1%) | 0.177 |
|  | Simple | 252 (91.6%) | 74 (91.4%) | 326 (91.6%) |  |
|  | Unmanageable | 0 (0.0%) | 1 (1.2%) | 1 (0.3%) |  |
| Adverse effects from ART | Never | 151 (54.9%) | 49 (60.5%) | 200 (56.2%) | 0.017 |
|  | Yes, and majors | 2 (0.7%) | 4 (4.9%) | 6 (1.7%) |  |
|  | Yes, but minors | 120 (43.6%) | 28 (34.6%) | 148 (41.6%) |  |
| ART treatment tolerance | No | 39 (14.2%) | 8 (9.9%) | 47 (13.2%) | 0.355 |
|  | Yes | 233 (84.7%) | 73 (90.1%) | 306 (86.0%) |  |
| Missed ART doses | Never | 109 (39.6%) | 64 (79.0%) | 173 (48.6%) | 0.0 |
|  | Regularly | 6 (2.2%) | 0 (0.0%) | 6 (1.7%) |  |
|  | Sometimes | 160 (58.2%) | 17 (21.0%) | 177 (49.7%) |  |
| Any ART change | Yes | 49 (17.8) | 37 (45.7) | 86 (24.2) | <0.001 |
| Reason for ART change | Effectiveness | 41 (14.9) | 12 (14.8) | 53 (14.9) | 1.000 |
|  | Tolerance | 23 (8.4) | 29 (35.8) | 52 (14.6) | <0.001 |
|  | Adherence | 7 (2.5) | 0 (0.0) | 7 (2.0) | 0.358 |
| Hospital visit frequency | Every 2–3 months | 168 (61.1) | 74 (91.4) | 242 (68.0) | < 0.001 |
|  | Every 6 months | 107 (38.9) | 7 (8.6) | 114 (32.0) |  |
| Partner aware of HIV status | I prefer not to say | 16 (5.8%) | 5 (6.2%) | 21 (5.9%) | 0.537 |
|  | No | 96 (34.9%) | 23 (28.4%) | 119 (33.4%) |  |
|  | Yes | 162 (58.9%) | 53 (65.4%) | 215 (60.4%) |  |
| Family aware of HIV status | I prefer not to say | 14 (5.1%) | 1 (1.2%) | 15 (4.2%) | 0.062 |
|  | No | 126 (45.8%) | 48 (59.3%) | 174 (48.9%) |  |
|  | Yes | 133 (48.4%) | 32 (39.5%) | 165 (46.3%) |  |
| Friends aware of HIV status | I prefer not to say | 15 (5.5%) | 3 (3.7%) | 18 (5.1%) | 0.573 |
|  | No | 223 (81.1%) | 70 (86.4%) | 293 (82.3%) |  |
|  | Yes | 36 (13.1%) | 8 (9.9%) | 44 (12.4%) |  |
