## Supplementary Table 2 for "“The injection gives freedom” - An exploration of long-acting injectable HIV treatment acceptance among patients seeking care in two Nairobi tertiary hospitals": S2 _ Table.docx

S2 Table. Advantages of LAI-ART among responders

| **Response** | **KNH n (%)** | **AKUH n (%)** | **Combined n (%)** | **p-value** |
| --- | --- | --- | --- | --- |
| Stop taking treatments every day | 160 (77.7) | 40 (78.4) | 200 (77.8) | 1.000 |
| Ensure effectiveness for a given period | 20 (9.7) | 20 (39.2) | 40 (15.6) | 0.000 |
| Be sure not to forget my medication | 106 (51.5) | 37 (72.5) | 143 (55.6) | 0.011 |
| Hide from surroundings/colleagues | 88 (42.7) | 29 (56.9) | 117 (45.5) | 0.097 |
| Forget the disease | 90 (43.7) | 5 (9.8) | 95 (37.0) | 0.000 |
| Not thinking about it daily | 123 (59.7) | 6 (11.8) | 129 (50.2) | 0.000 |
| Other | 7 (3.4) | 0 (0.0) | 7 (2.7) | 0.393 |
