## Supplementary Table 3 for "“The injection gives freedom” - An exploration of long-acting injectable HIV treatment acceptance among patients seeking care in two Nairobi tertiary hospitals": S3 _ Table.docx

S3 Table. Disadvantages of LAI-ART among responders

| **Response** | **KNH n (%)** | **AKUH n (%)** | **Combined n (%)** | **p-value** |
| --- | --- | --- | --- | --- |
| Losing freedom to stop ART | 1 (0.5) | 0 (0.0) | 1 (0.4) | 1.000 |
| Fear of side effects | 130 (63.1) | 45 (88.2) | 175 (68.1) | 0.001 |
| Fear of injections/pricks | 65 (31.6) | 7 (13.7) | 72 (28.0) | 0.018 |
| Fear of being taken for a guinea pig | 42 (20.4) | 4 (7.8) | 46 (17.9) | 0.059 |
