## Supplementary Table 4 for "“The injection gives freedom” - An exploration of long-acting injectable HIV treatment acceptance among patients seeking care in two Nairobi tertiary hospitals": S4 _ Table.docx

S4 Table. Preferences for Dosing

| **Characteristic** | **Category/Statistic** | **KNH n (%)** | **AKUH n (%)** | **Combined n (%)** | **p-value** |
| --- | --- | --- | --- | --- | --- |
| Preferred duration between injections | 1 month | 12 (5.8) | 1 (2.0) | 13 (5.1) | 0.017 |
|  | 2 months | 6 (2.9) | 21 (41.2) | 27 (10.5) |  |
|  | 3 months | 52 (25.2) | 18 (35.3) | 70 (27.2) |  |
|  | 6 months | 85 (41.3) | 7 (13.7) | 92 (35.8) |  |
